# The Association Between Platelet Transfusion and Mortality Rate Among Preterm Neonates in the Eastern Province, Saudi Arabia

**DOI:** 10.1101/2021.08.22.21262181

**Authors:** Fatimah Abdullah Alhamad, Ahlam Mohammed Hussien, Friyal Mubarak Alqahtani

## Abstract

Platelet transfusion is the main mode of management of thrombocytopenia. However, some studies link frequent and high-threshold platelet transfusions with an incremental increase of mortality rate.

**Aim:** This study aims to assess the association between the frequency and the threshold of platelet transfusions, with the mortality rate among preterm neonates.

**Methods:** A retrospective cohort study design was used. This study was conducted at maternity and children’s hospitals in Al-Ahasa, Saudi Arabia. The sample size includes 154 preterm neonates, included in the study by the use of the convenience sampling technique.

**Result:** There is a significant relationship found between the gestational age and the birth weight of the preterm neonates with the survival rates among both groups. In contrast, there is no significant relationship found between transfusion frequency, transfusion threshold, and the survival rates of the group which received platelet transfusion.

**Implications for Practice and Research:** The current study found that mortality is mainly associated with lower gestational ages, and not platelet transfusions. More studies are needed to fill the remaining gaps of knowledge, and to optimise platelet transfusion practices among preterm neonates.

## Background

Platelet transfusion is the main therapy for neonatal thrombocytopenia (platelet count < 150,000/μL), aside from directly treating the underlying cause.^1^ Thrombocytopenia is one of the most common hematologic problems faced by neonates, especially those admitted to neonatal intensive care units (NICU), and those with lower gestational ages.^2,3^ Moreover, around 22% - 35% of all neonates, and approximately 50% of ill preterm neonates (who are born alive before completing 37 weeks of gestation) suffer from thrombocytopenia.^4,5^ Theoretically, neonatal thrombocytopenia is correlated with a high risk of bleeding and mortality.^6^ Hence, platelet transfusions are needed to prevent the risk of haemorrhage and death.

Neonatal platelet transfusion thresholds vary according to clinical conditions. More liberal transfusions are considered by many neonatologists when facing cases of unstable preterm neonates, most neonatologists prefer restrictive transfusions for term neonates (who are born between 37 and 41 weeks of gestation).^7^ However, due to the recognised risks of platelet transfusions, and the frequent identification of bleeding among the disease conditions, rather than of low platelet count, restrictive platelet transfusions for preterm neonates is recommended.^1,8^

Theoretically, platelet transfusions at the high threshold (platelet < 50,000/μL) may reduce the risk of bleeding and mortality, though in practice this is not the case.^6^ There is a newly raised debate regarding the association between platelet transfusion frequency, threshold, and the elevation of mortality rates among preterm neonates. Through randomised control trials (RCT) research, it has been reported that the prophylactic high threshold platelet transfusion is significantly associated with a higher mortality rate, with major bleeding, or with both to a more extreme degree than with low threshold transfusions (platelet < 25,000/μL).^9^ Overall, there is uncertainty as to whether the elevation of neonatal mortality rate is associated with platelet transfusions, or with other factors, such as the severity of their illnesses.^10,11^

As a result, there are new approaches recently used as alternatives to platelet transfusion, such as the administration of thrombopoietic growth factors, and thrombopoiesis-stimulating agents, done in hopes of reducing the risks associated with platelet transfusions.^11,12^ However, the medical responses to those new approaches has not been well recognised.^4^ Furthermore, studies regarding platelet transfusion practices and risk factors among neonates are too few; as such, this study aims to assess the association between the frequency and threshold of platelet transfusions, with mortality rates among preterm neonates, focusing on the Maternity and Children’s Hospital (MCH) in Al-Ahasa, Saudi Arabia. It is assumed that the results of this study will add to the body of evidence regarding platelet transfusion practices among preterm neonates.

### Literature Review

Retrospective studies have assumed that platelet transfusions themselves, independently of disease conditions, could lead to harmful outcomes in neonates - however randomized control trials (RCT) that test these harmful consequences are not in and of themselves sufficient for a conclusion.^9^ There is a newly raised debate over the association between platelet transfusions and the elevation of mortality rates among preterm neonates.

Curley et al^9^conducted a multicentre RCT examination, aiming to assess the optimal prophylactic platelet transfusion threshold, with a total sample size of 660 preterm neonates. The study compared the death and major bleeding incidences between the group which received a low platelet transfusion threshold and the group which was treated with a high threshold transfusion. The RCT result was quite unexpected - it was concluded that the prophylactic high threshold transfusion is significantly associated with a higher mortality rate, with major bleeding, or with both – to a more extreme degree than that of low threshold transfusions among preterm neonates.^9^ Being a large multicentred randomized control trial study, with a notably large sample size, makes this a strong study. Likewise, a retrospective cohort study which was conducted in 2018 in Austria concluded that mortality rate is significantly increased as the number of platelets transfused increases (p < 0.05). Although that study involved a large sample size (371) of neonates, it was conducted on a single-centre fashion, and used a retrospective design. These factors can be considered as the study’s limitations.^12^ Furthermore, Kasap et al^10^ in their study in Turkey, found that the mortality rate among neonates who receive platelet transfusions was significantly higher than among those who did not receive platelet transfusions (p < 0.001). This study also used a retrospective design, though it used a larger sample size (395 patients) to overcome the limitation of retrospective design.

In contrast, several studies attributed the mortality rates to the underlying diseases, rather than to the transfusion frequency or threshold. One prospective multicentre cohort study concluded that the mortality among the neonates is correlated with disease severity, rather than the transfusion itself. However, although this study was multicentred, the sample size was small (n = 44), which makes it more difficult for researchers to find significant associations between platelet transfusions and mortality rates.^13^ Likewise, Dogra et al^14^ in their study in India, found that there is no association between the number of platelet transfusions and mortality. Yet, this study also had limitations in terms of its small sample size (n = 49), and additionally failed to perform medical follow-ups on the subjects involved.^14^ A retrospective cohort study conducted in the Kingdome of Saudi Arabia (KSA) by Alsafadi^15^ concurred with the previous study. It found that platelet transfusion was not correlated with mortality rates. It is important to note that this study was retrospective, and did not consider all of the factors that could be the leading causes of death.^15^

Overall, there is uncertainty as to whether or not the elevation of neonatal mortality rates is associated with platelet transfusions or with other factors, such as illness severity.^10,11^

Although the practice of platelet transfusions is common in NICUs, there is a wide variety of platelet transfusion practices and guidelines among neonatologists, relating to the involved threshold, and the risk and benefit of transfusion. According to the literature, frequent platelet transfusions among neonates are correlated with the elevation of mortality rate. This correlation was explained by many assumptions, such as the volume and speed of transfusion, and the pro-inflammatory feature of the platelets. In contrast, others believed that there is no association between platelet transfusions and death - they attributed the elevation of mortality rate to the severity of the underlying diseases themselves. Generally, there is uncertainty over platelet transfusion practices among neonates.

## Method

The field’s literature frequently assumed that frequent and high threshold platelet transfusions raise the mortality rates of preterm neonates.^9,12^ Therefore, the researchers of the current research conducted a retrospective cohort study in order to test this assumption, and to answer the research question.

The investigators measured the exposure to platelet transfusions and the survival status of the participants, thereby aiming to assess the association between the frequency and threshold of platelet transfusions, and the mortality rates among preterm neonates at the Maternity and Children’s Hospital (MCH) in Al-Ahasa, Saudi Arabia. This aim was fulfilled by covering four objectives: assessing the association between platelet transfusion frequency and mortality rates among preterm neonates, assessing the association between platelet transfusion thresholds and mortality rates among preterm neonates, assessing the association between mortality and the subjects’ demographic data, and assessing the association between mortality and the maternal characteristics.

A cohort design in this study represents two groups - one is of the preterm neonates who received platelet transfusions, and the other also is of preterm neonates who did not receive platelet transfusion therapy. Those two groups were compared against one another in terms of survival status.

To control extraneous variations and to safeguard internal validity, the current study selected the preterm neonates based on inclusion and exclusion criteria. The preterm neonates were selected based on the following inclusion criteria: the status of being preterm neonates (gestational age < 37 weeks) admitted to NICU within the period of January 2019 to December 2020; of being preterm neonates who received platelet transfusions, assigned to be the exposed group; and being preterm neonates who did not receive platelet transfusions, assigned to be unexposed group. The exclusion criteria were being preterm neonates with congenital malformation, genetic disorders, comorbid diseases, and the preterm neonates who were admitted from other health care institutions. The exclusion of those groups was due to the conditions themselves elevating the risk of mortality.^16,17^

As for the selected setting - the MCH at Al-Ahasa city in KSA; Al-Ahasa’s central referral hospital - the total number of admitted preterm neonates throughout 2019 and 2020 was 790. Thus, 60% of this number (474) was examined. Random sampling was completed through the use of a reliable website (https://www.dcode.fr/random-sampling). Out of this total number, only 77 received platelet transfusions. Therefore, every one of the neonates who received transfusions were chosen as the exposed group. In accordance with this, the same number of neonates with similar characteristics who did not receive platelet transfusions was selected as the unexposed group. Thus, the final number of the sample was 154 preterm neonates.

The obtained data were secondary data taken from the registry books and electronic systems of specific departments; namely, the Neonatal Intensive Care Unit (NICU), the blood bank department, haematology department, microbiology department, and the ultrasound department. The used data collection tool (Case Report Forms) was adopted and modified from Platelets for Neonatal Transfusion - Study 2 (PlaNeT – 2).^9^ To use the PlaNeT – 2 tool, permission was acquired by the authors. After some modifications, and after adding new items, the forms are merged into one form which contains five parts (demographic data, medical diagnosis, mother data, platelet transfusions information, and outcome) (see supplemental table 1). The face and content validity of the tool have been secured. Moreover, the reliability of the tool was tested. The Cronbach’s alpha was .733.

## The Result

Upon data completion and validation, the data was transported from the data collection sheet to the statistical software, IBM SPSS software package version 25.0, for analysis. Qualitative data were described using numbers and percentage. The significance of the obtained results was judged to be at the 5% level. The utilised tests were the Chi-squared test for categorical variables, which was used to compare the results between the two groups, as well as either Fisher’s exact test, or the Monte Carlo correction, the latter of which was used as a correction for the chi-squared test, when more than 20% of the cells showed an expected count less than 5.

The sample was divided into two groups of equal numbers (n = 77). The first group consisted of preterm neonates who were treated with platelet transfusions, whereas the second group included preterm neonates who did not receive platelet transfusions. The 77 preterm neonates in the transfusion group received a total of 296 platelet transfusions. Table (1) clarifies the sample characteristics. The most common admission diagnoses for the preterm neonates who received platelet transfusions were extremely preterm birth, extremely low birth weight, and respiratory distress (42.9%) - whereas the most common admission diagnoses among the group who did not receive platelet transfusion were preterm birth and respiratory distress (64.9%). Regarding the chief complaint, the most common chief complaint in both groups was of a grade I intraventricular haemorrhage (IVH). By exploring subjects’ mother’s data, the statistics showed that there was no medical history or antenatal complications among most mothers in both groups. Concerning the frequency of platelet transfusion, most of the subjects received platelet transfusions between 1 and 3 times (59.7%). Regarding the survival status across the two groups, the survival rate among the group who received platelet transfusion was more than half (61%), and the mortality was 39% - whereas for the other group the survival rate was 80.5%.

Concerning the relationship between survival status and demographic data of the study’s sample, there was a significant relationship between gestational age and birth weight of the preterm neonates with survival status in the group who received platelet transfusions; χ2 (MCp) = 10.120 (0.012), 11.565 (0.002), as well as with the group who did not receive platelet transfusions; χ2 (MCp) = 22.877 (< 0.001), 26.175 (< 0.001) respectively. Thus, as gestational age and birth weight decreases, the mortality rate increases. However, there was no significant relationship between the gender, the mode of delivery, and the number of gestations with survival status among the preterm neonates in both groups. The relationship between the medical diagnosis of the preterm neonates, their mothers’ data, and the survival status is represented on Table 2.

Concerning the relationship between the platelet transfusion frequency of the group who were treated with platelet transfusion and survival status, there was no significant relationship found (χ2 (MCp) 3.921 (0.398)). Moreover, the relationship between the pre-transfusion platelet count (the threshold) with survival status was studied. The statistical analysis showed that there was no significant relationship between the platelet transfusion threshold and survival status.

## Discussion

Platelet transfusion is used as a treatment or prophylactic therapy for preterm neonates with cases of thrombocytopenia. However, platelet transfusion is assumed to elevate the mortality rate among neonates. Hence, the current study tested this assumption retrospectively by using a retrospective cohort design. The current study examined 154 preterm neonates, of which 77 were assigned to be in the study group, and 77 were assigned to the control group.

Regarding the demographic and medical data of the selected sample, including neonates who either did or did not received platelet transfusions, the female gender was slightly more prevalent in the total sample. In contrast, Alsafadi,^15^ who studied in Saudi Arabia, and Kasap et al^10^ who conducted research in Turkey, reported that male gender was slightly more frequent in their respective samples. Their results were in agreement with the result of Peelen et al^18^ who reported that male gender is at higher risk of spontaneous premature delivery, and premature rupture of membrane (PROM) delivery than the female gender. The current study’s odd result in this regard may have been due to the fact that it was conducted on a one centre basis, and had a relatively small sample size.

Based on the sample collecting process, in which the group which received platelet transfusions was selected as the study group, extremely preterm neonates (GA < 28 weeks) were found to be the highest age group among the entire sample. The reason behind this is that the lower the gestational age, the lower the platelet count - which by extension necessitates platelet transfusions.^4^ Thereafter, the control group - which did not receive platelet transfusions - was specifically selected to share the same gestational ages (extremely preterm). This clearly explains why extremely preterm neonates represent the most dominant age group among the two groups.

Concerning the birth weight, it was found among the two groups to be varied. The most frequent birth weight (BW) of the group which received platelet transfusion was extremely low birth weight (BW< 1000 g), which matches with the findings of Curley et al^9^ whereas the most frequent birth weight of the group which did not receive platelet transfusions was a very low birth weight (BW<1500 g). This result is likely a consequence of having a high percentage of extremely preterm neonates.

The current study found that the most dominant diagnoses of subjects who received platelet transfusions were extremely preterm birth, extremely low birth weight, and respiratory distress - for those who did not receive platelet transfusions the most common diagnoses were preterm birth and respiratory distress. This finding is similar to the results reported by Tekleab et al^19^. This result is supported by the fact that the extremely preterm group is most often in need of platelet transfusions, as their platelet count tends to be low.^4^ For this reason, extremely preterm, extremely low birth weight, and respiratory distress diagnoses were the most dominant diagnoses among those who received platelet transfusion.

The most frequent chief complaint among both groups was that of intraventricular haemorrhage (IVH) grade I, which aligns with the results of Kasap et al.^10^ This finding could be attributed to the fact that IVH is associated with lower gestational age (GA < 32 weeks gestation),^20^ and the majority of the current study’s sample were extremely preterm neonates (GA < 28 weeks), followed by severely preterm neonates (GA 31-28 weeks).

The platelet transfusion details were explored in the current study. The frequency of platelet transfusion was investigated, and it was found that the most common number of transfusions was from 1 to 3 transfusions. This result was near to the results of Sparger et al,^21^ who reported that the mean (SD) of platelet transfusions among their study sample was 4.3. Concerning the platelet transfusion threshold, the category of platelets < 100.000/μL was found to be the most dominant threshold for platelet transfusion, which matches what was reported by Sparger et al.^21^ In contrast, a study conducted by Dogra et al^14^ found that platelet threshold between 20,000 and 50,000 μL was the most common threshold for platelet transfusions. To our knowledge, platelet transfusion thresholds vary worldwide based on institutions’ policies and procedures. Regardless, there is a lack of reliable evidence which guides clinical practices to identify the ideal thresholds for neonatal platelet transfusion.^5,22^

Neonatal transfusion triggers or thresholds vary according to clinical conditions, like gestational age at birth.^7^ The British Committee for Standards in Haematology’s 2016 guidelines, and the Dutch Guidelines Quality Council in 2018 set 25,000/μL as the threshold for prophylactic transfusion in stable neonates, and 50,000/μL in cases of bleeding, or of invasive procedures being necessary.^11^ As for another example, Sola-Visner and Bercovitz^23^ assumed that in the absence of good evidence, platelet transfusions for neonates should be done at a higher threshold, as neonates’ platelets are hypo-reactive. Indeed, the more liberal transfusions are considered necessary by many neonatologists for unstable preterm neonates, whereas they prefer restrictive transfusions for term neonates.^7^ Likewise, Jacquot et al^22^ reported that the risk of developing IVH forces many physicians to practice aggressive platelet transfusions (platelet count □ 100.000/μL) in high-risk patients. The last two suggestions justify why the transfusion threshold < 100.000/μL was the most dominant platelet transfusion practice in the current study.

Regarding the survival status of the two groups, the current study found that survival rates were higher than mortality rates among the two groups; either those who received platelet transfusions, or those who did not receive platelet transfusions. By investigating the demographic data of those who died, we could determine the causes of death as relating to the gestational age. The results show that there was a significant relationship between both gestational age and birth weight with survival rates in both groups. As the gestational age and birth weight decreases, the mortality rates increase. The results showed that 44.2% of those who received platelet transfusion and died were extremely preterm neonates (GA < 28 weeks). This result is identical to the results reported by Jang et al^24^ and Pammi^25^.

On one hand, the current study found that there was no significant relationship between transfusion frequency and survival rates. This result matches those of Alsafadi et al^15^ and Dogra et al^14^. Likewise, Sola-Visner et al^23^ assumed that the association between platelet transfusions and mortality rates mainly reflects the seriousness of the underlying conditions, or of extreme prematurity rather, than the transfusions themselves.^23^ On the other hand, Kasap et al^10^ found that the mortality rate was significantly higher among the group which received platelet transfusions compared to the group which did not receive platelet transfusions (p < 0.001). Additionally, Patel et al^13^ found a significant relationship between the transfusion frequency and the mortality rate - however, after considering the illness severity among those who died, the association between death and platelet transfusion became insignificant.

Concerning the relationship between the platelet counts before platelet transfusions (the threshold) with survival rates, there was no significant relationship found. This result is contrary to the findings of Curley et al.^9^ Namely, Curley et al^9^ found that a prophylactic high threshold transfusion was significantly associated with higher mortality rates when compared to low threshold transfusions among preterm neonates.

Though the current study did not find a relationship between platelet transfusions threshold or frequencies with mortality rates, we cannot fully reject the association between platelet transfusion and mortality, as many justifications may justify this hypothesis. There are many suggestions that explain the reason behind the harmful effects of the current platelet transfusion practices for neonates.^26^ One of those suggestions is that the volume and transfusion speed may lead to harm through hemodynamic effects.^26^ Traditionally, platelets are transfused for over 30 to 60 minutes, which leads to sudden a introduction of a large volume of bolus to a sick acidotic neonate with passive cerebral pressure circulation.^26^ Furthermore, it has been suggested that neonatal platelets are hyporeactive when compared to adults’ platelets, so it is unknown whether the transfusion of adult’s platelets to neonates induces micro-thrombosis formation or not. The last assumption is that platelets possess significant immunologic and inflammatory effects.^9,26,27^ Due to the fact that the exact causes developing adverse effects (such as death) of platelet transfusion has not been confirmed, restrictive platelet transfusions in preterm neonates are preferred over liberal transfusions.

## Recommendations

Based on the current study result, and considering the paucity of evidence regarding platelet transfusion among preterm neonates, it is recommended researchers conduct strong multi-hospital randomised control trials (RCT); this could contribute to a strong evidence-based practice, and could lead to the initiation of a clear guideline to be strictly followed. Furthermore, it is recommended that future studies use large sample sizes. Moreover, to our knowledge, only one study had been conducted in Saudi Arabia. Thus, it is recommended that more studies approach this topic in the KSA, and worldwide as well. Besides this, it is recommended that researchers consider all of the confounding variables in future studies, as this will aid the collection of valid results.

## Data Availability

The data is available with the researcher as well as on the medical records at the selected setting.

## Notes

### Competing Interest Statement

The authors have declared no competing interest.

### Funding Statement

There is no funding for this research

### Author Declarations

The approval for this study was obtained from ethical committee in the Standing Committee for Research Ethics on Living Creatures (SCRELC) Imam Abdulrahman bin Faisal University, Saudi Arabia. Then, an approval was taken from the Research Ethics Committee (REC) of Maternity and Children Hospital (MCH) at Alhasa city in Kingdome of Saudi Arabia.

