## Supplemental Material for "The Association Between Platelet Transfusion and Mortality Rate Among Preterm Neonates in the Eastern Province, Saudi Arabia"

**Table 1.**

***Data Collection Sheet***

| Demographic Data | |
| --- | --- |
| Log Number: | Gender: □ Male □ Female |
| Date of birth: □ / / 2020 □ / / 2019 | |
| Gestational Age (GA):  □ Late preterm (34-36 weeks) □ Moderate preterm (32-33 weeks)  □ Severe preterm(31-28 weeks) □ Extremely preterm ( GA<25 weeks) | |
| Birth Weight (BW):  □ Low birth weight (BW <2500 g) □Very low birth weight (BW<1500 g)  □ Extremely low birth weight (BW ≤1000 g) | |
| Mode of delivery:  □ Spontaneous Vaginal Delivery (SVD) □ Cesarean Section (C/S) □ Instrumental Delivery | |
| Baby blood group:  □ O^+^ □ O^-^ □ A^+^ □ A^-^ □ B^+^ □ B^-^ □ AB^+^ □ AB^-^ □ mixed field | |
| Number of gestations:  □ Single □ Twins □ Triplet □ Quadruplet □ Other ……. | |
| Medical Diagnosis | |
| Admission Diagnosis:  □ Preterm □ Respiratory distress (RD) □ Respiratory distress syndrome □ Low birth weight □ Intrauterine growth restriction □ Other……. | |
| Chief Complaint:  □ Intraventricular hemorrhage (IVH) □ Gram positive sepsis □ Gram negative sepsis  □ Necrotizing enterocolitis (NEC) □ Other………… | |
| Mother Data | |
| Medical diagnosis:  □ Non □ Diabetes milieus □ Hypertension □Hypothyroidism □ Other ……. | |
| Antenatal Complication:  □ Non □ Pre-eclampsia □ Premature rupture of membrane (PROM)  □Abruptio Placentae □ Breech presentation □ Emergency cesarean section □ Other ……. | |
| Mother Blood group:  □ O^+^ □ O^-^ □ A^+^ □ A^-^ □ B^+^ □ B^-^ □ AB^+^ □ AB^-^ | |
| Platelet (Plt) Transfusion Information | |
| Total number of platelet transfusion: | |
| □ Transfusion 1   - The age during transfusion: □ 1 – 10 □ 11 – 20 □ 21 – 30 - Plt unit ABO: □ O^+^ □ O^-^ □ A^+^ □ A^-^ □ B^+^ □ B^-^ □ AB^+^ □ AB^-^ - Plt count pre-transfusion: □ Plt <25.000/ μL □ Plt < 50.000/μL □ Plt < 100.000/μL   □ Plt < 150.000/μL □ Plt ˃150.000/μL | |
| □ Transfusion 2   - The age during transfusion: □ 1 – 10 □ 11 – 20 □ 21 – 30 - Plt unit ABO: □ O^+^ □ O^-^ □ A^+^ □ A^-^ □ B^+^ □ B^-^ □ AB^+^ □ AB^-^ - Plt count pre-transfusion: □ Plt <25.000/ μL □ Plt < 50.000/μL □ Plt < 100.000/μL   □ Plt < 150.000/μL □ Plt ˃150.000/μL ). | |
| □ Transfusion 3   - The age during transfusion: □ 1 – 10 □ 11 – 20 □ 21 – 30 - Plt unit ABO: □ O^+^ □ O^-^ □ A^+^ □ A^-^ □ B^+^ □ B^-^ □ AB^+^ □ AB^-^ - Plt count pre-transfusion: □ Plt <25.000/ μL □ Plt < 50.000/μL □ Plt < 100.000/μL   □ Plt < 150.000/μL □ Plt ˃150.000/μL | |
| □ Transfusion 4   - The age during transfusion: □ 1 – 10 □ 11 – 20 □ 21 – 30 - Plt unit ABO: □ O^+^ □ O^-^ □ A^+^ □ A^-^ □ B^+^ □ B^-^ □ AB^+^ □ AB^-^ - Plt count pre-transfusion: □ Plt <25.000/ μL □ Plt < 50.000/μL □ Plt < 100.000/μL   □ Plt < 150.000/μL □ Plt ˃150.000/μL | |
| □ Transfusion 5   - The age during transfusion: □ 1 – 10 □ 11 – 20 □ 21 – 30 - Plt unit ABO: □ O^+^ □ O^-^ □ A^+^ □ A^-^ □ B^+^ □ B^-^ □ AB^+^ □ AB^-^ - Plt count pre-transfusion: □ Plt <25.000/ μL □ Plt < 50.000/μL □ Plt < 100.000/μL   □ Plt < 150.000/μL □ Plt ˃150.000/μL | |
| □ Transfusion 6   - The age during transfusion: □ 1 – 10 □ 11 – 20 □ 21 – 30 - Plt unit ABO: □ O^+^ □ O^-^ □ A^+^ □ A^-^ □ B^+^ □ B^-^ □ AB^+^ □ AB^-^ - Plt count pre-transfusion: □ Plt <25.000/ μL □ Plt < 50.000/μL □ Plt < 100.000/μL   □ Plt < 150.000/μL □ Plt ˃150.000/μL | |
| □ Transfusion 7   - The age during transfusion: □ 1 – 10 □ 11 – 20 □ 21 – 30 - Plt unit ABO: □ O^+^ □ O^-^ □ A^+^ □ A^-^ □ B^+^ □ B^-^ □ AB^+^ □ AB^-^ - Plt count pre-transfusion: □ Plt <25.000/ μL □ Plt < 50.000/μL □ Plt < 100.000/μL   □ Plt < 150.000/μL □ Plt ˃150.000/μL | |
| □ Transfusion 8   - The age during transfusion: □ 1 – 10 □ 11 – 20 □ 21 – 30 - Plt unit ABO: □ O^+^ □ O^-^ □ A^+^ □ A^-^ □ B^+^ □ B^-^ □ AB^+^ □ AB^-^ - Plt count pre-transfusion: □ Plt <25.000/ μL □ Plt < 50.000/μL □ Plt < 100.000/μL   □ Plt < 150.000/μL □ Plt ˃150.000/μL | |
| □ Transfusion 9   - The age during transfusion: □ 1 – 10 □ 11 – 20 □ 21 – 30 - Plt unit ABO: □ O^+^ □ O^-^ □ A^+^ □ A^-^ □ B^+^ □ B^-^ □ AB^+^ □ AB^-^ - Plt count pre-transfusion: □ Plt <25.000/ μL □ Plt < 50.000/μL □ Plt < 100.000/μL   □ Plt < 150.000/μL □ Plt ˃150.000/μL | |
| □ Transfusion 10   - The age during transfusion: □ 1 – 10 □ 11 – 20 □ 21 – 30 - Plt unit ABO: □ O^+^ □ O^-^ □ A^+^ □ A^-^ □ B^+^ □ B^-^ □ AB^+^ □ AB^-^ - Plt count pre-transfusion: □ Plt <25.000/ μL □ Plt < 50.000/μL □ Plt < 100.000/μL   □ Plt < 150.000/μL □ Plt ˃150.000/μL | |
| □ Transfusion 11   - The age during transfusion: □ 1 – 10 □ 11 – 20 □ 21 – 30 - Plt unit ABO: □ O^+^ □ O^-^ □ A^+^ □ A^-^ □ B^+^ □ B^-^ □ AB^+^ □ AB^-^ - Plt count pre-transfusion: □ Plt <25.000/ μL □ Plt < 50.000/μL □ Plt < 100.000/μL   □ Plt < 150.000/μL □ Plt ˃150.000/μL | |
| □ Transfusion 12   - The age during transfusion: □ 1 – 10 □ 11 – 20 □ 21 – 30 - Plt unit ABO: □ O^+^ □ O^-^ □ A^+^ □ A^-^ □ B^+^ □ B^-^ □ AB^+^ □ AB^-^ - Plt count pre-transfusion: □ Plt <25.000/ μL □ Plt < 50.000/μL □ Plt < 100.000/μL   □ Plt < 150.000/μL □ Plt ˃150.000/μL | |
| □ Transfusion 13   - The age during transfusion: □ 1 – 10 □ 11 – 20 □ 21 – 30 - Plt unit ABO: □ O^+^ □ O^-^ □ A^+^ □ A^-^ □ B^+^ □ B^-^ □ AB^+^ □ AB^-^ - Plt count pre-transfusion: □ Plt <25.000/ μL □ Plt < 50.000/μL □ Plt < 100.000/μL   □ Plt < 150.000/μL □ Plt ˃150.000/μL | |
| □ Transfusion 14   - The age during transfusion: □ 1 – 10 □ 11 – 20 □ 21 – 30 - Plt unit ABO: □ O^+^ □ O^-^ □ A^+^ □ A^-^ □ B^+^ □ B^-^ □ AB^+^ □ AB^-^ - Plt count pre-transfusion: □ Plt <25.000/ μL □ Plt < 50.000/μL □ Plt < 100.000/μL   □ Plt < 150.000/μL □ Plt ˃150.000/μL | |
| □ Transfusion 15   - The age during transfusion: □ 1 – 10 □ 11 – 20 □ 21 – 30 - Plt unit ABO: □ O^+^ □ O^-^ □ A^+^ □ A^-^ □ B^+^ □ B^-^ □ AB^+^ □ AB^-^ - Plt count pre-transfusion: □ Plt <25.000/ μL □ Plt < 50.000/μL □ Plt < 100.000/μL   □ Plt < 150.000/μL □ Plt ˃150.000/μL | |
| Outcome | |
| Final outcome: □ Survive □ Died, if yes, the date of death …………. | |
